# Central Nervous System Complications of Invasive Group A Streptococcal Infection in Children: A UK National Cohort Study, 2022-2025

**DOI:** 10.64898/2026.11.27.26369129

**Authors:** Erva Nur Cinar, Penelope J. Teoh, Victoria Holdstock, Conor John O’Donovan, Anu Goenka, Anya Stephenson, Barbara Chow, Christo Tsilifis, Marieke Emonts, James E.G. Charlesworth, James E. Burns, Fiona Shackley, Thomas C. Williams, Louisa Pollock, Eiman Hamid, George Lawson, Nuria Sanchez Clemente, Matthew Laundy, James Stewart, Michael J Carter, James Weitz, Ana Cockburn, Benedict Michael, Shivaram Avula, Rachel M. Roberts, Alison Kent, Andrew Riordan, Stephen T. J. Ray

**Affiliations:** Department of Paediatric Infectious Diseases and Immunology, Evelina London Children’s Hospital, Guy’s and St Thomas’ NHS Foundation Trust, London SE1 7EH, United Kingdom; Department of Clinical Microbiology, Great Ormond Street Hospital for Children NHS Trust, London WC1N 3JH, United Kingdom; Department of Paediatric Infectious Diseases and Immunology, Royal Hospital for Children, NHS Greater Glasgow and Clyde, Glasgow G51 4TF, United Kingdom; Department of Paediatric Infectious Diseases and Immunology, Bristol Royal Hospital for Children, University Hospitals Bristol NHS Foundation Trust, Bristol BS2 8BJ, United Kingdom; Bristol Vaccine Centre, Bristol Medical School, University of Bristol, Bristol BS8 1TD, United Kingdom; Department of Paediatric Infectious Diseases and Immunology, St Mary’s Hospital, Imperial College Healthcare NHS Trust, London W2 1NY, United Kingdom; Department of Paediatrics, London North West University Healthcare NHS Trust, Harrow HA1 3UJ, United Kingdom; Department of Paediatric Immunology, Infectious Diseases and HSCT, Great North Children’s Hospital, Newcastle upon Tyne NE1 4LP, United Kingdom; Translational and Clinical Research Institute, Newcastle University, Newcastle upon Tyne NE1 7RU, United Kingdom; Department of Paediatric Infectious Diseases and Immunology, Oxford Children’s Hospital, Oxford University Hospitals NHS Foundation Trust, Oxford OX3 9DU, United Kingdom; Centre for Human Genetics, University of Oxford, Oxford OX3 7BN, United Kingdom; Department of Laboratory Medicine and Pathobiology, University of Toronto, Toronto M5S 3K9, Canada; Department of Paediatric Infectious Diseases and Immunology, Sheffield Children’s Hospital, Sheffield Children’s NHS Foundation Trust, Sheffield S10 2TH, United Kingdom; Department of Child Life and Health, University of Edinburgh, Edinburgh EH16 4TJ, United Kingdom; School of Infection and Immunity, University of Glasgow, Glasgow G12 8TA, United Kingdom; Department of Paediatric Infectious Diseases and Immunology, Great Ormond Street Hospital, London WC1N 3JH, United Kingdom; Department of Paediatric Infectious Diseases, St George’s Hospital, St. George’s University Hospital NHS Foundation Trust, London SW17 0QT, United Kingdom; Department of Infection, St George’s Hospital, St George’s University Hospitals NHS Foundation Trust, London SW17 0QT, United Kingdom; Department of Medical Paediatrics, Royal Aberdeen Children’s Hospital, Aberdeen AB25 2ZG, United Kingdom; Paediatric Critical Care Unit, John Radcliffe Hospital, Oxford University Hospitals NHS Foundation Trust, Oxford OX3 9DU, United Kingdom; Victoria Hospital Kirkcaldy, NHS Fife, Kirkcaldy, KY2 5AH, United Kingdom; Department of Clinical Infection, Microbiology and Immunology, Institute of Infection, Veterinary and Ecological Sciences, University of Liverpool, Liverpool, United Kingdom; Department of Neurology, The Walton Centre NHS Foundation Trust, Liverpool L9 7LJ, United Kingdom; Department of Radiology, Alder Hey Children’s NHS Foundation Trust, Liverpool L14 5AB, United Kingdom; Department of Neuroradiology, Oxford University Hospitals NHS Foundation Trust, Oxford OX3 9DU, United Kingdom; Department of Paediatric Infectious Diseases, Alder Hey Children’s NHS Foundation Trust, Liverpool L12 2AP, United Kingdom; Oxford Vaccine Group, Department of Paediatrics, University of Oxford, Oxford OX3 7LE, United Kingdom; The Brain Infection and Inflammation Group, University of Liverpool, Liverpool L9 7AL, United Kingdom; Malawi–Liverpool Wellcome Trust Clinical Research Programme, P.O. Box 30096, Blantyre 3, Malawi; Blantyre Malaria Project, Kamuzu University of Health Sciences, P.O. Box 32256, Blantyre 3, Malawi; Wilson Lab, Weill Institute for Neurosciences, University of California, San Francisco, San Francisco, CA 94158, United States

## Abstract

**Background:** Central Nervous System (CNS) invasive Group A Streptococcal infection (iGAS) in children is uncommon but severe; the clinical spectrum during the iGAS resurgence post-COVID pandemic is poorly described.

**Methods:** We conducted a UK multicenter national cohort study of children aged <18 years admitted from January 2022 to March 2025 with CNS complications of iGAS disease. Clinician-reported data were reviewed by a panel of pediatric infectious diseases specialists and neuroradiologists.

**Results:** Forty-eight children were included; median age was 57 months, 29/48 (60%) were male. Antecedent neurological risk factors were recorded in 28/48 (58%), most commonly otitis media in 10/48 (21%), mastoiditis in 10/48 (21%) and sinus infection in 6/48 (13%). The predominant CNS phenotypes were subdural empyema in 16/48 (33%), meningitis in 13/48 (27%), and intracranial thrombosis in 9/48 (19%). Diagnostic confirmation was by culture alone in 34/48 (71%) and PCR alone in 5/48 (10%). Neuroimaging showed suppurative intracranial disease in 33/48 (69%) and venous sinus or cerebral venous thrombosis in 18/48 (38%). 22/48 (46%) children required intensive care and 37/48 (77%) underwent surgery including Ear, Nose, and Throat (ENT) procedures in 17/48 (35%) and neurosurgical procedures in 25/48 (52%). At late outcome assessment, 6/47 (13%) had died and 15/47 (32%) had neurological sequelae.

**Conclusions:** In this largest European national cohort, CNS iGAS was associated with death or neurological sequelae in 21/47 (45%) children with known late outcomes. Mortality was greater than meningococcal or pneumococcal meningitis adverse outcome estimates in UK children. Long-term neurodevelopmental follow-up is essential.

**Summary of the article’s main points:** In this UK national cohort of 48 children, invasive group A streptococcal CNS complications were heterogeneous and severe, frequently requiring intensive care and surgery, with death or neurological sequelae in nearly half of children at late outcome assessment.

## Background

Group A *Streptococcus* (GAS, *Streptococcus pyogenes*) causes a wide spectrum of disease in children, ranging from pharyngitis and cellulitis to invasive infections. Invasive GAS (iGAS) infection is conventionally defined by detection of GAS from a normally sterile site, whereas disease severity is determined by the clinical manifestations, including shock and organ dysfunction [1]. Severe GAS disease may be associated with sepsis, streptococcal toxic shock syndrome, and death. Central nervous system (CNS) involvement is rare but clinically important. It can cause rapidly progressive neurological deterioration necessitating intensive care, prolonged antimicrobial therapy and surgical source control, with associated high case fatality rates and long-term neurological disability [2–4].

The epidemiology of pediatric invasive Group A Streptococcus (iGAS) changed markedly after the relaxation of SARS-CoV-2 pandemic restrictions. From late 2022, several European and North American countries reported an increase of pediatric iGAS, including fatal cases. This coincided with high respiratory virus circulation and increased GAS transmission [5,6]. In the United Kingdom (UK), this period was also associated with a resurgence of both non-invasive (often scarlet fever) and laboratory-confirmed invasive GAS disease, with attention drawn to *emm*1 and the M1UK lineage in national and international surveillance [7]. Although these epidemiological signals have been widely reported, the clinical spectrum of CNS complications during this period remains poorly characterized.

Existing pediatric literature on GAS CNS disease is limited. Most are case reports or small series focusing on GAS meningitis, brain abscess or suppurative intracranial infections. A US surveillance study described intracranial GAS infections in children and a recent systematic review focused on GAS meningitis [2,4]. However, robust national-level European data describing the clinical outcomes and broader CNS phenotypes observed with pediatric iGAS, are scarce.

We therefore established a national pediatric cohort of children with iGAS and CNS complications. In this study, we describe the cohort demographics, clinical presentation, iGAS and neurological risk factors, microbiology, CNS syndromes, imaging findings, antimicrobial and surgical management, and early and late outcomes.

## Methods

### Study Design and Setting

We performed a multicenter national retrospective cohort study of children with CNS complications of iGAS infection in the UK. Eligible cases were children aged <18 years admitted between January 2022 and March 2025. Cases were identified through a UK national clinician-reporting process established by an expert pediatric infectious diseases group for CNS complications of iGAS. Clinicians were invited to submit anonymized case data through a standardized online reporting portal.

### Case Definitions

Hospitalized children were eligible if they had acute-onset CNS disease and laboratory-confirmed or clinically suspected iGAS infection considered by the referring pediatric infectious diseases clinician to be the cause of the CNS presentation. Detection of GAS from a normally sterile site by culture or molecular testing provided microbiological confirmation of invasive infection. Clinically suspected cases were also included when GAS was detected from a non-sterile site in a child and was considered the likely cause of the CNS presentation by an expert in pediatric infectious diseases. Final inclusion and CNS phenotype classification were agreed by pediatric infectious diseases specialists, with neuroradiology input where imaging findings were relevant, over the course of three months. Clinically phenotype-based analyses retained the predefined diagnostic categories assigned by the referring clinicians; neuroimaging findings were summarized separately. Children were excluded if the presentation was Pediatric Autoimmune Neuropsychiatric Disorders Associated with Streptococcal Infection, mastoiditis alone or pre-septal cellulitis alone (Detailed CNS phenotype definitions, inclusion and exclusion criteria are presented in Supplementary Appendix 1A).

### Data Collection

Data were collected using a standardized electronic case report form covering demographics, medical history, risk factors, clinical presentation, investigations, microbiology, treatment, intensive care, surgical interventions, and outcomes (complete case report form is provided in Supplementary Appendix 1B). Antecedent risk factors were classified as non-neurological when they reflected systemic or extracranial infection outside CNS, and as neurological when they represented potential predisposing factors for CNS infection, including ENT/parameningeal structures, neurosurgical or anatomic factors, or head trauma.

Imaging and electroencephalography (EEG) reports were captured as anonymized free-text (complete case report form is provided in Supplementary Appendix 1B). Cases were reviewed by a panel of experts in pediatric infectious diseases and two neuroradiologists.

### Outcomes

Early outcomes referred to status at the end of the acute treating-hospital admission. Early death referred to death during that admission, early neurological sequelae referred to survival with neurological sequelae at discharge or transfer, and early adverse outcome was the composite of these two outcomes.

Late outcomes reflected the most recent recorded clinical status at the time of case submission and were classified using modified Glasgow Outcome Scale (GOS) categories [8]: full recovery or return to baseline (score 5), minor neurological sequelae (score 4), moderate neurological sequelae (score 3), severe neurological sequelae (score 2), or death (score 1).

### Statistical Analysis

Descriptive analyses included categorical variables (counts and percentages) and continuous variables (median and interquartile range).

Data were analyzed using descriptive statistics (counts, percentages, medians, and interquartile ranges) and group comparison tests with Mann-Whitney U or Kruskal-Wallis tests for continuous or ordinal variables and two-sided Fisher exact tests for 2×2 categorical variables. Given the small cohort size and descriptive aims, no multivariable modeling was performed. Between-group comparisons were exploratory, and P values were two-sided and were not adjusted for multiple comparisons. Findings were interpreted alongside effect estimates, confidence intervals and subgroup sizes and were considered hypothesis-generating.

### Governance and Ethics

The study was deemed to be a quality improvement project, was registered locally as such, and had a UK Health Research Authority (HRA) ethical approval waiver. Only anonymized data was submitted by responding clinicians. Individual patient consent and referring-site-specific ethical approval were not required for clinician-entered anonymized data collection.

## Results

### Cohort Characteristics and Clinical Presentation

Forty-eight children with central nervous system (CNS) complications of invasive Group A Streptococcus (iGAS) were included. The median age was 57 months (IQR 20.8-113.3). 29/48 (60%) children were male. Most children were tertiary referrals (transferred from secondary care general pediatric hospital setting): 34/48 (71%); 14/48 (29%) were admitted directly from community or outpatient services. (Supplementary Table 1). Clinical characteristics, investigations, management, and outcomes by CNS phenotype are summarized in Table 1.

**Table 1.**
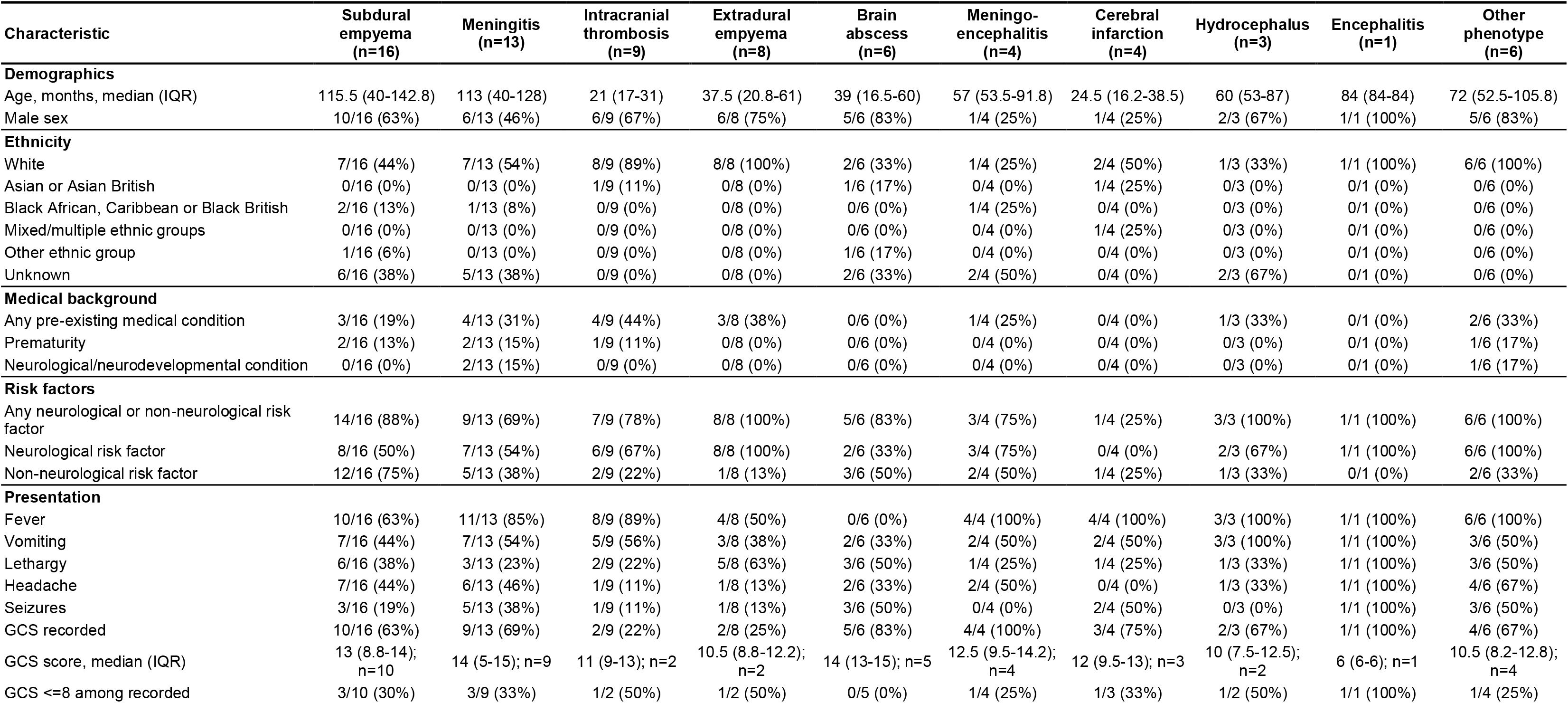
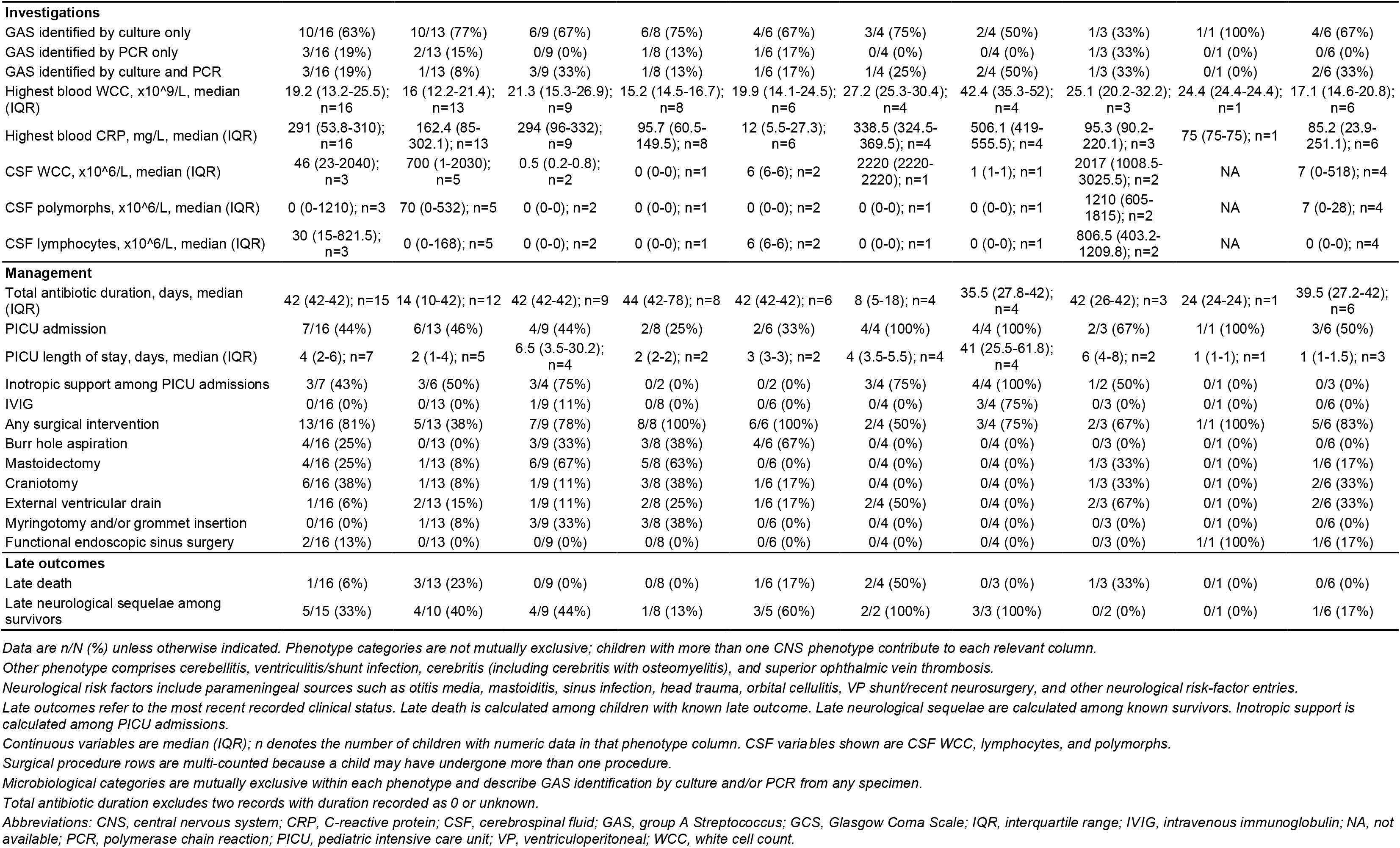
Clinical characteristics and outcomes by CNS phenotype.

9/48 (19%) children had a pre-existing medical condition. Prematurity was the most frequent background condition, in 3/48 (6%) children. Among 34 children with available data, 33/34 (97%) were fully immunized for age (Supplementary Table 1).

Antecedent non-invasive non-neurological risk factors for iGAS were identified in 21/48 (44%) children; most commonly pharyngitis or tonsillitis in 12/48 (25%), followed by varicella-zoster virus (VZV) infection in 3/48 (6%) and skin or soft tissue infection in 2/48 (4%). Antecedent neurological risk factors were recorded in 28/48 (58%) and most commonly included otitis media in 10/48 (21%), mastoiditis in 10/48 (21%), sinus infection in 6/48 (13%) and head trauma in 5/48 (10%) (Supplementary Table 1).

The median duration of non-neurological risk-factor symptoms was 8 days (IQR 5.8-14) and 3 days for neurological risk factors (IQR 2-7). Presenting symptoms had a median duration of 4 days (IQR 2-7).

Neurological symptoms were the most common presenting symptom group in 41/48 (85%). The most frequent individual symptoms were fever in 32/48 (67%), vomiting in 23/48 (48%), lethargy in 20/48 (42%), headache in 16/48 (33%), seizures in 14/48 (29%), and drowsiness in 14/48 (29%). Glasgow Coma Score (GCS) was recorded in 29 children, with a median score of 12; 22/29 (76%) had a GCS <15, including 8/29 (28%) with a GCS ≤8 (Supplementary Table 1).

### CNS Phenotype, Microbiology and Investigations

The most common CNS complications from iGAS were subdural empyema in 16/48 (33%), meningitis in 13/48 (27%), intracranial thrombosis in 9/48 (19%), extradural empyema in 8/48 (17%), and brain abscess in 6/48 (13%). Meningoencephalitis was reported in 4/48 (8%) children (Table 1; Supplementary Table 2). GAS was isolated exclusively from non-sterile sites in four children (4/48, 8.3%).

GAS was identified by culture alone in 34/48 (71%), PCR alone in 5/48 (10%), and both in 9/48 (19%) cases. Culture sample types most commonly included pus in 22/48 (46%), blood in 13/48 (27%), throat swab in 7/48 (15%), and CSF in 6/48 (13%). PCR sample types most commonly included pus in 5/48 (10%), cerebrospinal fluid (CSF) in 4/48 (8%) and blood in 3/48 (6%) (Supplementary Table 3).

CSF was successfully obtained in 16/48 (33%) children. GAS was detected in CSF by at least one microbiological method in 8/16 (50%); culture positive in 6, GAS-specific CSF PCR positive in 3 and 16S PCR positive in 1.

Median CSF white cell count (WCC) was 6 x 10^6/L^ (IQR 1-700), median CSF glucose was 3.1 mmol/L (IQR 2.8-3.4) and median CSF protein was 0.29 g/L (IQR 0.24-0.72); this varied by CNS phenotype. Blood inflammatory markers were available for all 48 children. The median highest white cell count was 19.47 x 10^9/L^ (IQR 14.1-25.7) and the median highest neutrophil count was 13.79 x 10^9/L^ (IQR 9.2-22). The median highest platelet count was 520 x 10^9/L^ (IQR 393.3-641) and the median highest C-reactive protein was 159.2 mg/L (IQR 30.7-304). Exploratory comparisons showed higher WCC with cerebral infarction, higher neutrophil counts and CRP with cerebral infarction or meningoencephalitis, lower platelet counts with meningitis, and lower CRP with brain abscess (Table 1 and Supplementary Table 4).

Antimicrobial susceptibility details were available for 11/43 (26%) culture-positive cases. All 11 cases were sensitive to penicillin. *emm* typing was available in 28/48 (58%) and within this group, *emm*1.0 was the most common type in 16/28 (57%). (Supplementary Table 5).

Neuroimaging was performed in most children: MRI head in 41/48 (85%), CT head in 44/48 (92%), and follow-up MRI head in 37/48 (77%). Baseline MRI reports were available in 39 children and were abnormal in all. CT reports were available in 41 children and were abnormal in 40/41 (98%). In the child whose CT was reported as normal, an earlier MRI had demonstrated a right frontal extradural abscess with adjacent leptomeningeal enhancement and following MRI showed resolution of the collection with residual mild leptomeningeal enhancement. Suppurative intracranial disease was common in 33/48 (69%) children, with subdural empyema or collection in 20/48 (42%), extradural or epidural empyema or collection in 11/48 (23%), and brain abscess in 6/48 (13%). Vascular complications were also frequent, including venous sinus or cerebral venous thrombosis in 18/48 (38%), cerebral infarction or ischemia in 11/48 (23%), and hemorrhage in 11/48 (23%). Other common imaging features included meningeal enhancement or imaging features of meningitis in 29/48 (60%), cerebritis, cerebellitis, or encephalitis in 14/48 (29%), hydrocephalus in 7/48 (15%), mass effect, midline shift, herniation, or effacement in 26/48 (54%), and cerebral or vasogenic edema in 18/48 (38%). Parameningeal source foci were frequently identified, particularly mastoiditis or middle ear disease, and sinusitis or paranasal sinus disease; each recorded in 20/48 (42%) children. Follow-up MRI most commonly showed improvement with residual abnormality in 22/37 (59%), while 5/37 (14%) showed progression or a new abnormality. These findings included progressive white matter changes with a new fluid collection, hemispheric ischemia with mass effect, increased cerebellar swelling with ventricular enlargement, worsening meningoencephalitis with new ischemic changes, and an infected frontal soft-tissue collection communicating with the frontal sinus.

EEG was performed in 9/48 (19%) children and was abnormal in all 9.

### Management, Disease Course, and Outcomes

Prior antimicrobial status was known for 42/48 (88%) children; 39/42 (93%) had received antimicrobials before admission or transfer. All 34 children referred or transferred to tertiary care had received antimicrobials before transfer. Among the 13 direct admissions, prior antimicrobial status was known for eight children, of whom 5/8 (63%) had received antimicrobials before admission. Status was unknown for the remaining five direct admissions and the child admitted from outpatient services. The most common prior antimicrobial regimen included third-generation intravenous (IV) cephalosporins in 33/39 (85%). Empiric and definitive antimicrobial therapy mostly included ceftriaxone in 27/48 (56%) and 35/48 (73%), respectively. Clindamycin was included in empiric regimens in 10/48 (21%) children and definitive regimens in 7/48 (15%), with 3 children receiving it during both empiric and definitive treatment. Median total antimicrobial duration was 42 days (Figure 2).

**Figure 1.**
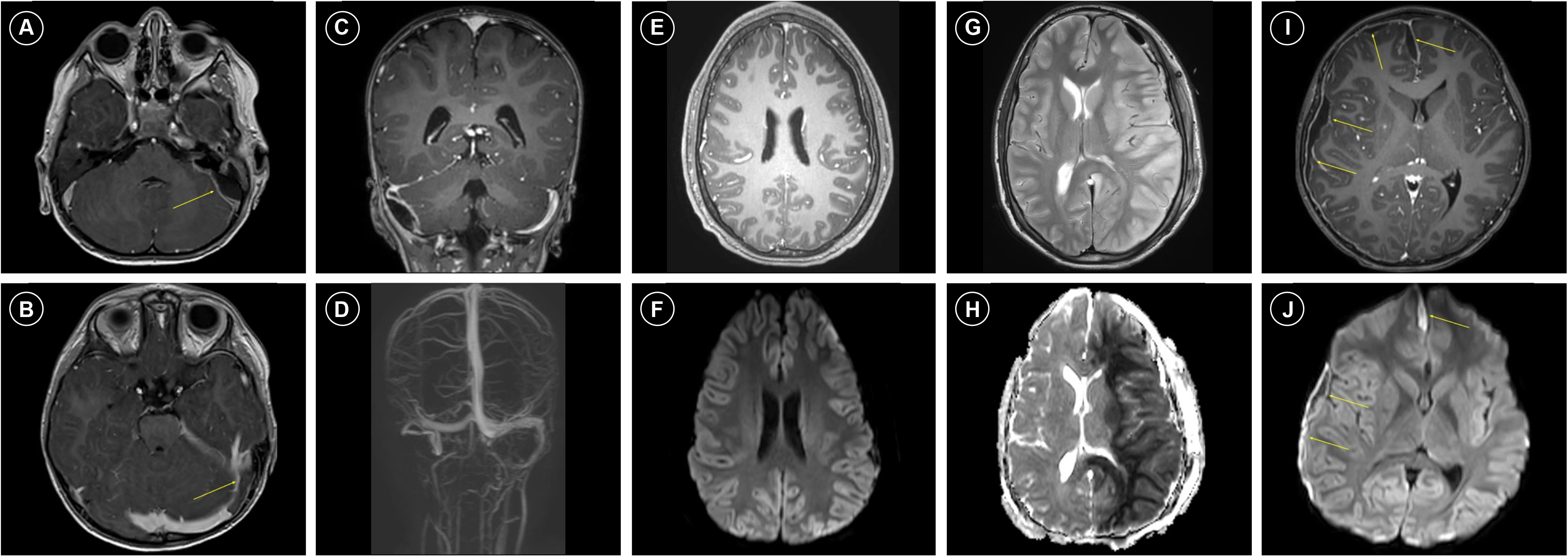
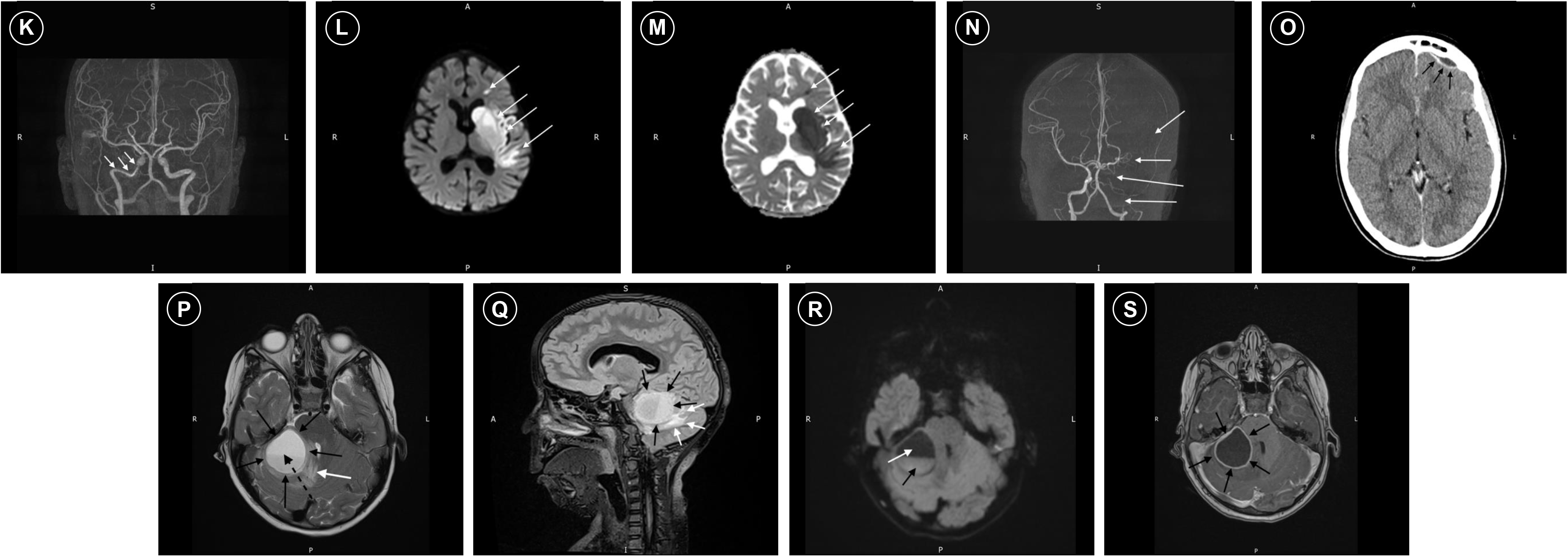
Neuroimaging findings in iGAS CNS disease. Representative MRI findings showing suppurative and vascular complications of central nervous system invasive group A Streptococcus. (A, B) Left posterior fossa extradural collection related to mastoiditis with adjacent transverse/sigmoid sinus and internal jugular vein thrombosis. (C, D) Right-sided complicated mastoiditis with epidural fluid collection and right sigmoid sinus thrombosis. (E, F) Left subdural collection with dural enhancement, cortical diffusion restriction consistent with cerebrit is, and suspected venous thrombus. (G, H) Postoperative images showing marked left cerebral edema and restricted diffusion consistent with ischemia. (I, J) Right subdural empyema with mass effect and a small adjacent cerebral abscess. *Abbreviations: CNS, central nervous system; iGAS, invasive group A streptococcal disease; MRI, magnetic resonance imaging*. Representative vascular and posterior fossa imaging findings. (K) Time-of-flight angiography showing narrowing of the right internal carot id artery. (L, M) Diffusion-weighted imaging and apparent diffusion coefficient maps showing acute left middle cerebral artery territory infarction. (N) Time-of-flight angiography showing absent normal signal in the left internal carotid and middle cerebral arteries, consistent with occlusion. (O) Post-contrast CT showing a left frontal extra-axial collection consistent with empyema. (P-S) Right cerebellar thick-walled cyst ic lesion with surrounding edema, atypical diffusion appearances, and avid rim enhancement on post-contrast T1 imaging. *Abbreviations: ADC, apparent diffusion coefficient; CNS, central nervous system; CT, computed tomography; DWI, diffusion-weighted imaging; ICA, internal carotid artery; iGAS, invasive group A streptococcal disease; MCA, middle cerebral artery; MRI, magnetic resonance imaging; TOF, time-of-flight*.

**Figure 2.**
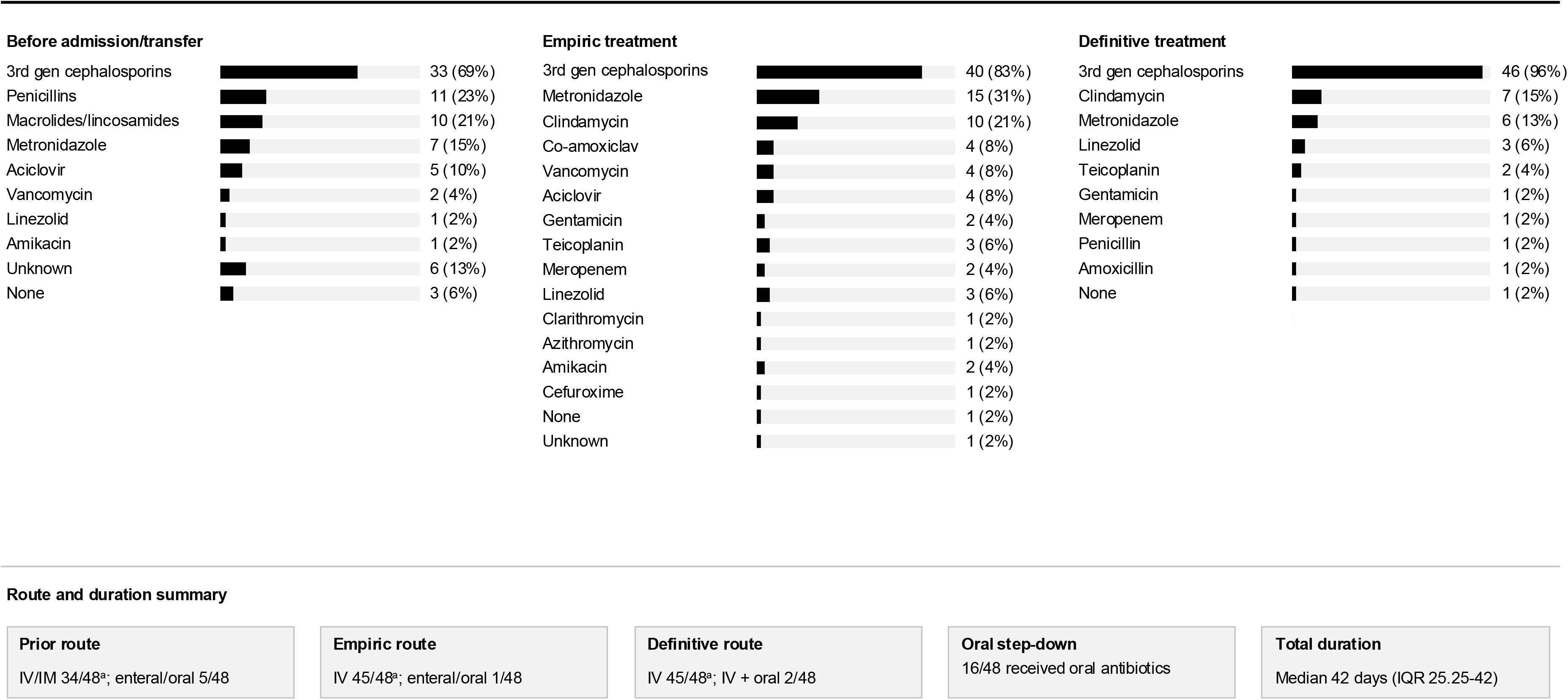
Antimicrobial treatment in iGASCNS disease. Third-generation cephalosporins (ceftriaxone, cefotaxime and ceftazidime) are grouped across all phases, counting each child once per phase. Other agents are shown individually, except prior macrolides/lincosamides. Gentamicin was recorded in one child with concomitant endocarditis as part of a treatment course involving multiple agents, rather than as sole definitive therapy. Prior macrolide/lincosamide treatment includes azithromycin, clindamycin, clarithromycin, and erythromycin, as defined in theanalysis. ^ᵃ^ Gaps reflect patients with no antimicrobial route recorded for that phase: prior route, none 3 and unknown 6; empiric route, none 1 and unknown 1; definitive route, not recorded 1. Abbreviations: CNS, central nervous system; iGAS, invasive group A streptococcal disease; IM, intramuscular; IQR, interquartile range; IV, intravenous.

Intravenous immunoglobulin (IVIG) was given to 3/48 (6%) children. Two with clinician-reported streptococcal toxic shock syndrome and one with severe septic shock and multiorgan failure. All three required PICU admission and inotropic support. All 3 children who received IVIG also received empiric clindamycin; none had clindamycin recorded as part of definitive treatment.

Surgical intervention was performed in 37/48 (77%) children. The most frequent procedure was burr hole aspiration in 11/48 (23%). 8/48 (17%) children required further surgery, including 4 who required further neurosurgical intervention (Supplementary Table 6). PICU admission was required in 22/48 (46%) children. Among those admitted to PICU, median PICU duration was 3 days (IQR 2-9), 18/22 (82%) required invasive respiratory support and 12/22 (55%) required inotropic support (Supplementary Table 6). Intensive care requirements, adjunctive treatments, and selected surgical procedures by CNS phenotype are summarized in Table 1.

Venous sinus thrombosis was diagnosed in 15/48 (31%) children. Anticoagulation was given to 14/15 (93%) children with thrombosis (median duration of anticoagulation 14 weeks (IQR 12-25.5)).

At the end of the acute treating-hospital admission, 5/48 (10%) children had died and 13/48 (27%) survived with neurological sequelae. One of these 13 children subsequently died after transfer back to the referring hospital. Three additional children initially recorded as having no neurological sequelae were classified in the minor-sequelae category at late outcome assessment. Analysis of late outcomes included 47 children with known late outcome status. Death or neurological sequelae at late outcome assessment occurred in 21/47 (45%), including 6/47 (13%) deaths and neurological sequelae in 15/47 (32%) children (Figure 3). Late outcomes by CNS phenotype are presented in Table 1.

**Figure 3.**
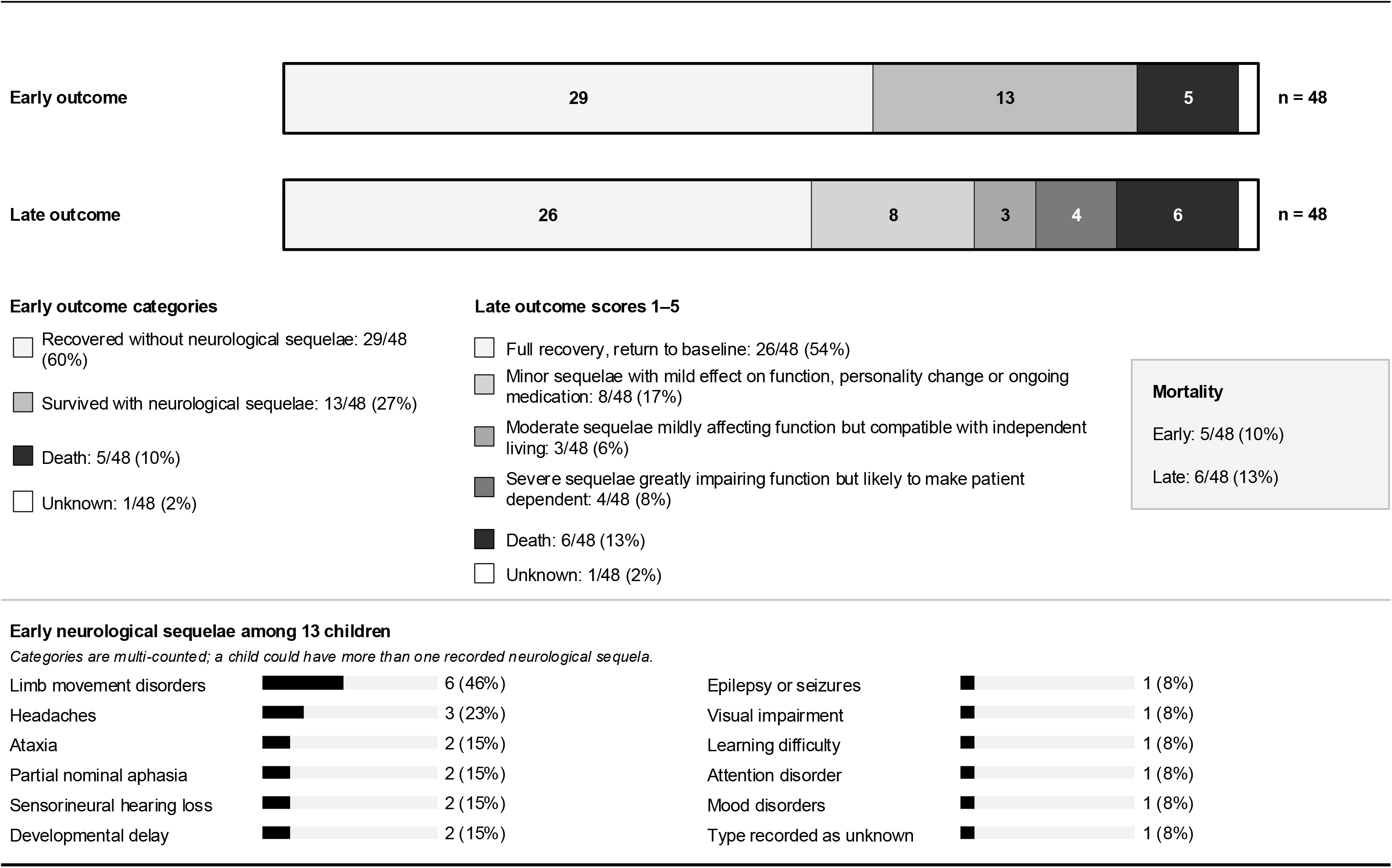
Early and late outcomes in iGAS CNS disease *Early outcomes: end of treating-hospital admission. Late outcomes: most recent assessment. Early adverse outcome: death or survival with neurological sequelae. Abbreviations: CNS, central nervous system; iGAS, invasive group A streptococcal disease*.

Death or neurological sequelae were more frequent among children admitted to PICU than not (14/21 (67%) vs 7/26 (27%); OR 5.43, 95% CI 1.55-19.0; P = 0.009). Children aged 5-9 years had lower odds of death or neurological sequelae at late outcome assessment than children in other age groups (2/12 (17%) vs 19/35 (54%); OR 0.17, 95% CI 0.03-0.88; P = 0.042). (Supplementary figure 1A).

All 4 children with meningoencephalitis died or had neurological sequelae at late outcome assessment (P = 0.034). Among children with cerebral infarction and known outcome status, all 3 had early neurological sequelae (P = 0.018), death or neurological sequelae at late outcome assessment (3/3 (100%); P = 0.082), and early adverse outcome (3/3 (100%); P = 0.050. Other phenotypes, including brain abscess, meningitis, subdural empyema, and intracranial thrombosis, showed variable outcome frequencies without significant evidence of association. (Supplementary Table 2; Supplementary Figure 1B).

## Discussion

In this UK national pediatric cohort from the 2022-2025 iGAS resurgence period, CNS complications of iGAS were rare but severe, with a variety of clinical phenotypes extending beyond meningitis. Nearly half of children required PICU admission and more than three quarters underwent at least one surgical procedure. Death or neurological sequelae affected over one third of children at the end of the acute treating-hospital admission. Subsequent follow-up captured an additional death following transfer to referring local hospital and minor sequelae in three children initially recorded as recovered without neurological sequelae. These findings suggest that assessment at discharge or transfer may not fully capture the subsequent burden of disease and support structured neurodevelopmental follow-up. The overall burden of disease suggests that CNS iGAS is a condition with high mortality which requires early multidisciplinary recognition, microbiological diagnosis, neuroimaging, antimicrobial treatment and where indicated, surgical source control.

Published intracranial GAS and GAS meningitis cohorts report mortality around 16-17% and neurological sequelae in over one quarter of survivors [2–4]. Late mortality in our cohort (13%) was greater than pediatric estimates: 5.3% for meningococcal and 9.9% for pneumococcal meningitis in France [9]. Pneumococcal meningitis mortality was 10.7% at 30 days among children aged <5 years in England and Wales (2000-2016) [10], and 9.9% in UK and Republic of Ireland childhood surveillance (2020-2022) [11]. UKHSA reported 2% mortality for meningococcal serogroup B meningitis alone and 5% with concomitant septicemia among children aged <5 years in England (2015-2021) [12]. Our cohort also adds UK data across CNS complications beyond meningitis.

The clinical pattern in this cohort supports prior reports highlighting the need for a low threshold for considering CNS complications in children with suspected or confirmed iGAS who have persistent fever, neurological deterioration, reduced consciousness, seizures, headache, vomiting, or focal neurological signs, particularly in the context of recent ENT or parameningeal infection and anatomical risk factors. Otitis media, mastoiditis, and sinus infection were common antecedent risk factors, consistent with previous GAS meningitis and intracranial GAS reports [2,4,13]. However, fever was not recorded in 16/48 (33%) children and neurological symptoms were not recorded in 7/48 (15%); presentations could also be non-specific and non-toxic, including lethargy or headache.

CSF sampling was achieved in only one third of children in our cohort, likely reflecting in part the source of infection originating outside of the CNS, the practical difficulty of lumbar puncture in children with intracranial collections, raised intracranial pressure, clinical instability, or urgent surgical priorities. Diagnosis therefore often depended on culture or PCR from blood, pus, or other clinically relevant samples. This supports a pragmatic diagnostic approach combining early neuroimaging with microbiological sampling from CSF when safe, and from sterile-site or source-control specimens when CSF is not feasible and keeping a low threshold for imaging, particularly in sinogenic iGAS with ongoing fevers and evolving neurological symptoms. Capturing CNS GAS cases in routine surveillance may also be challenging when GAS is detected only at non-sterile sites. In addition, our cohort demonstrated a high frequency of venous sinus thrombosis, making early consideration of vascular imaging important, especially when intracranial extension or thrombosis is suspected.

Most cases were culture-confirmed, all isolates with susceptibility data were penicillin-susceptible, and *emm*1.0 was the most frequent *emm* type among typed isolates. This is consistent with previous intracranial GAS and GAS meningitis surveillance in which *emm*1 was prominent [2,3]. It also aligns with UKHSA surveillance during the 2022-2023 iGAS surge, when *emm*1.0 was the most common invasive type overall and accounted for 65% of typed pediatric iGAS referrals, and with other post-pandemic UK pediatric iGAS phenotypes, including GAS pneumonia with parapneumonic effusion, in which *emm*1 predominated among typed isolates [14,15].

PICU admission, meningoencephalitis, and cerebral infarction were markers of more severe disease. Early adverse outcome also varied by age band, with the lowest proportion observed among children aged 5-9 years: 1/12 (8%). Mortality was numerically higher among infants aged <1 year, but this was based on one death among only three infants and was not statistically significant.

Most children had received antimicrobials before admission or transfer, definitive therapy was usually intravenous and beta-lactam-based. Adjunctive clindamycin and IVIG were used in a minority of cases, suggesting the management of CNS complications of iGAS, is distinct from classic toxin-mediated iGAS syndromes, although treatment indications were not consistently captured. In the absence of randomized pediatric data to guide adjunctive therapy or surgical timing, management requires coordinated input from infectious diseases, neurology, neurosurgery, radiology, and intensive care teams [16,17].

This study has several strengths. It is, to our knowledge, the first national UK pediatric cohort focused on CNS complications of iGAS during the post-2022 resurgence period and captures a broader phenotype than meningitis alone. Cases were systematically reported through a specialist pediatric infectious diseases network using a structured dataset, with information on presentation, microbiology, imaging, antimicrobial therapy, surgery, PICU care and outcomes.

The study also has limitations. Case ascertainment depended on clinician reporting and is like to have favored more severe, tertiary-referred or specialist-managed cases. Clinician-assigned CNS phenotypes did not always capture all subtle abnormalities reported on neuroimaging. These discrepancies may reflect subtle radiological findings that were not labeled as separate clinical diagnoses and may have influenced phenotype frequencies and phenotype-specific outcome comparisons. The cohort size is modest, limiting statistical power and preventing multivariable modeling. Due to the pragmatic data collection, several variables had missing, unknown or conditionally not-applicable data. Follow-up duration was not standardized, which may have been influenced by disease severity, potentially affecting the ascertainment of late neurological sequelae. CSF was not obtained in most children, further limiting direct comparison of CSF profiles across the cohort.

Overall, CNS complications of pediatric iGAS were associated with frequent PICU admission, vascular complications, need for surgical source control, substantial mortality, and neurological morbidity in survivors. Children with confirmed or suspected iGAS and evolving neurological features should have early CNS-penetrating antimicrobial therapy, urgent neuroimaging including vascular imaging where appropriate, microbiological sampling (including from CSF when possible and/or from sterile/surgical sites), early coordinated multidisciplinary management and structured neurodevelopmental long-term follow-up. Larger international collaborative studies with standardized follow-up and microbiological lineage data are urgently needed to better define risk factors, optimize treatment strategies, characterize long-term outcomes, and identify sequalae that may manifest after discharge.

## Data Availability

Requests for access to anonymised study data may be made to the corresponding author. Access is subject to the relevant governance approvals.

## Author contributions

E.N.C. developed the electronic case report form (Methodology), collected study data (Investigation), performed the statistical analyses (Formal analysis), coordinated the study and communication with contributors (Project administration), and drafted the manuscript (Writing).

A.K., A.R., and S.T.J.R. supported developing the research questions and study aims (Conceptualization) and designing the study methodology, including defining the inclusion and exclusion criteria (Methodology) and designing the case report form. They made substantial contributions to data analysis and interpretation of the results (Formal analysis) and provided study oversight and mentorship (Supervision).

S.A. and R.M.R. reviewed and interpreted the neuroimaging findings and selected images for inclusion in the manuscript (Investigation; Visualization).

P.J.T., V.H., C.J.O.D., A.G., A.S., B.C., C.T., M.E., J.E.G.C., J.E.B., F.S., T.C.W., L.P., E.H., G.L., N.S.C., M.L., J.S., M.J.C., J.W., A.C., and B.M. contributed case data from their respective centers (Investigation).

All authors contributed to critical review and revision of the manuscript (Writing, review and editing), approved the final version, and agreed to be accountable for all aspects of the work.

## Conflict of interest

STJR is supported by a UK Brain Research UK Project Grant, Thrasher child health early career award, Academy of Medical Sciences starter grant, and European Society of Paediatric Infectious Diseases Society diagnostic grant; is a Centre for Human Genetics Well Institute Olink-48 grant prize winner and Early Career Research committee member, both at The University of Oxford and sits on the Encephalitis International Scientific Committee

COD and AG acknowledge support from Spencer Dayman Meningitis Research, National Institute for Health and Care Research (NIHR ACF-2022-25-003), and NIHR Health Protection Research Focus Award in Vaccines and Immunisation at University of Bristol (NIHR207421), The views expressed are those of the author(s) and not necessarily those of the NIHR or the UK Department of Health and Social Care.

LP has received research funding from Pfizer Inc for a research topic unrelated to this manuscript.

## Funding resource

None

