## Supplementary materials for "Central Nervous System Complications of Invasive Group A Streptococcal Infection in Children: A UK National Cohort Study, 2022-2025"

Supplementary appendices, tables, and figure

** Required field in the original electronic survey.*

Table of contents


| Section | Contents |
|---|---|
| Supplementary Appendix 1A | Eligibility and phenotype-specific diagnostic criteria |
| Supplementary Appendix 1B | Case report form used for data collection |
| Supplementary Appendix 2 | Supplementary tables and figure |
| Supplementary Table 1 | Patient characteristics and presentation |
| Supplementary Table 2 | Early and late outcomes by CNS phenotype |
| Supplementary Table 3 | GAS detection by specimen source and method |
| Supplementary Table 4 | Blood and CSF markers by CNS phenotype |
| Supplementary Table 5 | emm typing results |
| Supplementary Table 6 | Clinical management and interventions |
| Supplementary Figure 1 | Factors associated with late outcomes |

Supplementary Appendix 1A. Eligibility and phenotype-specific diagnostic criteria

**Inclusion criteria**

☐ Child aged <18 years.

☐ Acute-onset central nervous system disease (e.g., meningitis, encephalitis, intracranial collection, venous sinus thrombosis, or orbital cellulitis).

☐ Laboratory-confirmed invasive group A streptococcal infection or suspected infection (e.g., high antistreptolysin O titre).

☐ Group A streptococcal infection implicated by the referring clinician as a possible cause of the acute-onset central nervous system disease.

**Exclusion criteria**

☐ A diagnosis of pediatric autoimmune neuropsychiatric disorder associated with streptococcal infection (PANDAS).

☐ A diagnosis of mastoiditis or preseptal cellulitis only.

### Phenotype-specific diagnostic criteria

| Phenotype | Criteria |
|---|---|
| **Meningitis** | Microbiologically confirmed OR suspected iGAS<br>**AND**<br>Child with symptoms consistent with meningitis* and:<br>1. CSF leukocyte count >4 × $10^6$ cells/L; OR<br>2. CSF leukocyte count ≤4 × $10^6$ cells/L AND one of:<br>a. Meningitis confirmed by MRI by a radiologist<br>b. Abnormal CSF biochemistry (protein >0.5 g/L and/or CSF:blood glucose ratio <0.5)<br>c. Immunocompromised (low CD4 count, high HIV viral load count [>1000 RNA copies/mL], or clinical evidence of advanced HIV disease) |
| **Encephalitis** | Microbiologically confirmed OR suspected iGAS<br>**AND**<br>Altered consciousness for >24 hours (lethargy, irritability, or change in personality) with two or more of:<br>1. Fever or history of fever (≥38°C) during the current illness<br>2. Seizures and/or focal neurological signs (with evidence of brain parenchymal involvement)<br>3. CSF pleocytosis (>4 × $10^6$ cells/L)<br>4. EEG suggesting encephalitis<br>5. Neuroimaging (MRI) suggestive of encephalitis |
| **Meningoencephalitis** | Microbiologically confirmed OR suspected iGAS<br>**AND**<br>Patients who fulfil the criteria for both meningitis and encephalitis. |
| **Subdural empyema** | Microbiologically confirmed OR suspected iGAS<br>**AND**<br>Clinical syndrome consistent with subdural empyema and confirmation with radiological evidence. |
| **Extradural empyema** | Microbiologically confirmed OR suspected iGAS<br>**AND**<br>Clinical syndrome consistent with extradural empyema and confirmation with radiological evidence. |
| **Intracranial thrombosis** | Microbiologically confirmed OR suspected iGAS<br>**AND**<br>Clinical syndrome consistent with intracranial thrombosis and confirmation with radiological evidence. |
| **Brain abscess** | Microbiologically confirmed OR suspected iGAS<br>**AND**<br>Clinical syndrome consistent with brain abscess and confirmation with radiological evidence. |
| **Hydrocephalus** | Microbiologically confirmed OR suspected iGAS<br>**AND**<br>Clinical syndrome consistent with hydrocephalus and confirmation with radiological evidence. |

*Abbreviations: ASO, antistreptolysin O; CNS, central nervous system; CSF, cerebrospinal fluid; EEG, electroencephalogram; GAS, group A Streptococcus; iGAS, invasive group A streptococcal disease; MRI, magnetic resonance imaging; PANDAS, pediatric autoimmune neuropsychiatric disorders associated with streptococcal infections.*

Supplementary Appendix 1B. Case report form used for data collection

**Instructions**

*Complete one form per patient. Do not enter identifiable patient information. Select all applicable responses where the electronic form permitted multiple selections. Questions shown after “if yes” or “if other” were displayed conditionally.*

**Details of hospital site and clinician**

**Hospital/site where patient was seen** *

________________________________

**Patient source** *

☐ Direct admission ☐ Tertiary referral/transfer ☐ Outpatient

**Date of admission/transfer** *

________________________________

**Referring clinician's full name** *

________________________________

**Referring clinician's email address** *

________________________________

**Institution/affiliation** *

________________________________

**Study participant reference number** *

*Enter the first two letters of the institution followed by the participant number (e.g., OX001). Do not enter a hospital or NHS number.*

________________________________

### Clinical inclusion criteria

**CNS complication phenotype as defined by the referring clinician ***

*See Supplementary Appendix 1A for the eligibility and phenotype-specific diagnostic criteria.*

☐ Meningitis ☐ Meningoencephalitis ☐ Encephalitis

☐ Extradural empyema ☐ Subdural empyema ☐ Brain abscess

☐ Intracranial thrombosis ☐ Hydrocephalus ☐ Cerebral infarction

☐ Other

**If other, please specify ***

______________________________

### Demographics

**Sex at birth ***

☐ Female ☐ Male ☐ Other

**If other, please specify ***

______________________________

**Patient age in months at presentation to your center (e.g., 2 years 10 months = 34 months) ***

______________________________

**Ethnicity ***

☐ White ☐ Mixed or multiple ethnic groups

☐ Asian or Asian British ☐ Black African, Caribbean, or Black British

☐ Other ethnic group ☐ Unknown

### Comorbidities and background

**Does the patient have any pre-existing medical conditions? ***

☐ Yes ☐ No ☐ Unknown

**If yes, please specify ***

☐ Prematurity

☐ Respiratory disease

☐ Neurological disease

☐ Gastrointestinal disease

☐ Kidney disease

☐ Genetic conditions

☐ Hematological/oncological disease

☐ Diabetes or other endocrinological diseases

☐ Other

☐ Immunosuppression/immunodeficiency

☐ Cardiac disease

☐ Neurodevelopmental conditions

☐ Liver disease

☐ Chronic infections

☐ Metabolic disease

**Please give further details of the patient's comorbidities ***

____________________________________________

**Please define the immunosuppression/immunodeficiency and give further details ***

____________________________________________

**Was the patient receiving immunosuppressive or immunomodulatory treatment before presentation? ***

☐ Yes ☐ No ☐ Unknown

**If yes, list the medications ***

____________________________________________

**Immunization status ***

☐ Fully immunized for age ☐ Incomplete ☐ Unknown

### Clinical presentation

**Did the patient have any of the following non-neurological (systemic) risk factors before presentation? ***

☐ Pharyngitis/tonsillitis ☐ Skin/soft tissue infection ☐ Dental procedures

☐ Varicella ☐ None of the above ☐ Other

**If other, please specify ***

______________________________________________________________

**Duration of non-neurological (systemic) symptoms before presentation (total number of days) ***

______________________________________________________________

**Did the patient have any of the following neurological risk factors before presentation? ***

☐ Head trauma ☐ Deep brain stimulation

☐ Ventriculoperitoneal shunt or another indwelling intracranial device

☐ CSF leak ☐ Meningocele ☐ Sinus infection

☐ Otitis media ☐ Mastoiditis ☐ Cochlear implant

☐ Orbital cellulitis ☐ Recent neurosurgery

☐ Recent epidural/lumbar puncture

☐ None of the above ☐ Other

**If recent neurosurgery, specify the surgical procedure ***

______________________________________________________________

**If recent neurosurgery, give the date of the surgical intervention ***

______________________________________________________________

**If other, please specify ***

______________________________________________________________

**Duration of neurological risk-factor symptoms before presentation (total number of days) ***

______________________________________________________________

**Presenting symptoms and signs ***

☐ Lethargy ☐ Seizures ☐ Coma

☐ Drowsiness ☐ Earache ☐ Headache

☐ Coryza ☐ Sore throat ☐ Fever

☐ Cough ☐ Difficulty breathing ☐ Rash

☐ Vomiting ☐ Diarrhea ☐ Muscle aches

☐ Photophobia ☐ Neck stiffness or pain ☐ Bulging fontanelle

☐ Hypotonia ☐ Hearing loss

☐ Focal neurological deficits (e.g., cranial nerve palsy, visual or speech disturbance, hemiparesis, or hemiplegia)

☐ Other

**If other, please specify ***

______________________________

**Duration of presenting symptoms before presentation (total number of days) ***

______________________________

**Which neurological-status measures were available at admission? ***

☐ AVPU ☐ Glasgow Coma Scale ☐ Neither available

**Glasgow Coma Scale score on admission ***

______________________________

**AVPU score on admission ***

☐ Alert ☐ Verbal ☐ Pain ☐ Unresponsive

**Admission diagnosis/impression ***

______________________________

**Evidence of group A Streptococcus and investigations**

### Blood tests

**Highest white blood cell count recorded ***

______________________________

**Highest neutrophil count recorded ***

______________________________

**Highest platelet count recorded ***

---

**Highest C-reactive protein recorded ***

---

**Antistreptolysin O titre result (enter N/A if unavailable) ***

---

### Microbiology

**How was group A Streptococcus identified in this patient? ***

☐ Positive culture for group A Streptococcus ☐ Positive PCR for group A Streptococcus

☐ Other

**If other, please specify ***

---

**If positive culture, specify the sample type tested ***

☐ Blood ☐ Throat ☐ Skin

☐ Pus ☐ Other

**If other, please give further details ***

---

**If positive PCR, specify the sample type tested ***

---

**If iGAS was identified by culture, were any antibiotic-resistant or intermediate susceptibilities detected? Please provide details. ***

---

**Was GAS emm typing performed? ***

☐ Yes ☐ No ☐ Unknown

**Typing result ***

---

**Was another bacterial or viral coinfection identified? ***

☐ Yes ☐ No ☐ Unknown

**Details of bacterial or viral coinfection, including confirmation method and results ***

---

**Was a cerebrospinal fluid sample obtained? ***

☐ Yes ☐ No ☐ Unknown

### Cerebrospinal fluid results

*Record the most abnormal CSF results, excluding traumatic taps.*

**Was the CSF culture positive for GAS? ***

☐ Yes ☐ No ☐ Unknown ☐ Not done

**Was broad-range 16S rRNA PCR from CSF positive for GAS? ***

☐ Yes ☐ No ☐ Unknown ☐ Not done

**Was GAS-specific PCR from CSF positive? ***

☐ Yes ☐ No ☐ Unknown ☐ Not done

**CSF total white blood cell count ***

---

**CSF lymphocyte count ***

---

**CSF polymorph count ***

---

**CSF red blood cell count ***

---

**CSF glucose ***

---

**Plasma glucose ***

---

**CSF protein ***

---

**Other CSF results**

---

### Neuroimaging and electroencephalography

**Did the patient undergo head MRI? ***

☐ Yes ☐ No ☐ Unknown

**If yes, provide the date and anonymized report of the initial MRI ***

---

**Did the patient undergo follow-up head MRI? ***

☐ Yes ☐ No ☐ Unknown

**If yes, provide the date and anonymized report of each follow-up MRI ***

---

**Did the patient undergo head CT? ***

☐ Yes ☐ No ☐ Unknown

**If yes, provide the date and anonymized report of each CT scan ***

---

**Did the patient undergo electroencephalography? ***

☐ Yes ☐ No ☐ Unknown

**If yes, EEG result ***

*Ensure that any free-text information is anonymized.*

☐ Normal ☐ Abnormal ☐ Unknown

**Details of other CNS imaging**

*Ensure that the information is anonymized.*

---

**Final diagnosis following investigations ***

---

### Management and progress

#### Antimicrobial therapy

**Were any of the following antibiotics given before transfer to your center? ***

☐ Third-generation cephalosporin

☐ Vancomycin ☐ Linezolid ☐ Carbapenem

☐ Other ☐ Unknown

**If other, please specify ***

______________________________________________________________________________

**Route of administration ***

☐ Intravenous ☐ Intramuscular ☐ Enteral ☐ Unknown

**Were any of the following antibiotics given in the community before admission to your center? ***

☐ Phenoxymethylpenicillin ☐ Amoxicillin ☐ Co-amoxiclav

☐ Azithromycin ☐ Erythromycin ☐ Clarithromycin

☐ Other ☐ Unknown

**If other, please specify ***

______________________________________________________________________________

**Route of administration ***

☐ Intravenous ☐ Intramuscular ☐ Enteral ☐ Unknown

**Empiric antibiotics on or following admission ***

☐ Phenoxymethylpenicillin ☐ Amoxicillin ☐ Co-amoxiclav

☐ Azithromycin ☐ Erythromycin ☐ Clarithromycin

☐ Clindamycin ☐ Cefalexin ☐ Ceftriaxone

☐ Cefotaxime ☐ Gentamicin ☐ Teicoplanin

☐ Vancomycin ☐ Meropenem ☐ Linezolid

☐ Unknown ☐ Other

**If other, please specify ***

______________________________________________________________________________

**Route of administration ***

☐ Intravenous ☐ Intramuscular ☐ Enteral ☐ Unknown

**Main antibiotic used for definitive treatment ***

☐ Ceftriaxone ☐ Cefotaxime ☐ Gentamicin

☐ Teicoplanin ☐ Vancomycin ☐ Meropenem

☐ Linezolid ☐ Unknown ☐ Other

**If other, please specify ***

______________________________________________

**Route of administration ***

☐ Intravenous ☐ Intramuscular ☐ Enteral ☐ Unknown

**Were oral antibiotics given following intravenous antibiotic treatment? ***

☐ Yes ☐ No ☐ Unknown

**Oral antibiotic used ***

☐ Phenoxymethylpenicillin ☐ Amoxicillin ☐ Co-amoxiclav

☐ Azithromycin ☐ Erythromycin ☐ Clarithromycin

☐ Clindamycin ☐ Cefalexin ☐ Teicoplanin

☐ Vancomycin ☐ Linezolid ☐ Unknown

☐ Other

**If other, please specify ***

______________________________________________

**Reason for oral antibiotics ***

______________________________________________

**Total duration of oral and intravenous/intramuscular antibiotic treatment (days) ***

______________________________________________

### Adjunctive treatment and complications

**Was intravenous immunoglobulin administered? ***

☐ Yes ☐ No ☐ Unknown

**Dose of intravenous immunoglobulin ***

______________________________________________

**Was a further dose of intravenous immunoglobulin administered? ***

☐ Yes ☐ No ☐ Unknown

**Dose of further intravenous immunoglobulin ***

---

**Intracranial complications ***

☐ Meningitis ☐ Meningoencephalitis ☐ Encephalitis

☐ Extradural empyema ☐ Subdural empyema ☐ Brain abscess

☐ Thrombosis ☐ Hydrocephalus ☐ Cerebral infarction

☐ Cerebromalacia

☐ No intracranial complication

☐ Other

**If other, please provide details ***

---

**Systemic complications ***

☐ Respiratory failure ☐ Disseminated intravascular coagulation

☐ Streptococcal toxic shock syndrome ☐ No systemic complications

☐ Other

**If other, please provide details ***

---

**Was venous sinus thrombosis diagnosed? ***

☐ Yes ☐ No ☐ Unknown

**If yes, was anticoagulation given? ***

☐ Yes ☐ No ☐ Unknown

**Duration of anticoagulation treatment (weeks) ***

---

### Intensive care and surgery

**Was the patient admitted to pediatric intensive care? ***

☐ Yes ☐ No ☐ Unknown

**Duration of pediatric intensive care stay (days) ***

---

**Was respiratory support required? ***

☐ Yes ☐ No ☐ Unknown

**Highest level of respiratory support ***

☐ Invasive ☐ Noninvasive ☐ Unknown

**Was inotropic support required? ***

☐ Yes ☐ No ☐ Unknown

**Did the patient undergo any surgical intervention? ***

☐ Yes ☐ No ☐ Unknown

**Type of surgical intervention ***

☐ Burr-hole aspiration ☐ Mastoidectomy ☐ Craniotomy

☐ External ventricular drain ☐ Other ☐ Unknown

**If other, please specify ***

______________________________________________________________________

**Did the patient undergo any further surgical intervention? ***

☐ Yes ☐ No ☐ Unknown

**Type of further surgical intervention ***

☐ Burr-hole aspiration ☐ Mastoidectomy ☐ Craniotomy

☐ External ventricular drain ☐ Other ☐ Unknown

**If other, please specify ***

______________________________________________________________________

### Outcome and follow-up

**Outcome ***

☐ Death ☐ Recovery with neurological sequelae

☐ Recovery without neurological sequelae ☐ Unknown

**If neurological sequelae were present, specify all that applied ***

☐ Sensorineural hearing loss ☐ Developmental delay

☐ Epilepsy/seizures

☐ Limb movement disorder (e.g., hemiplegia, diplegia, quadriplegia, paraplegia, or monoplegia)

☐ Visual impairment ☐ Learning difficulty

☐ Cranial nerve palsy ☐ Partial nominal aphasia

☐ Spatial-orientation impairment ☐ Memory deficit

☐ Headache ☐ Attention disorder

☐ Mood disorder ☐ Other

**If other, please provide details ***

______________________________________________

**Discharge destination ***

☐ Home ☐ Transferred back to referring center

☐ Rehabilitation/physiotherapy ☐ Deceased

☐ Unknown

**Date of discharge/death/transfer ***

______________________________________________

**Outcome at last follow-up ***

☐ 5 - Full recovery; returned to baseline

☐ 4 - Minor sequelae with mild effect on function, personality change, or ongoing medication

☐ 3 - Moderate sequelae mildly affecting function but compatible with independent living

☐ 2 - Severe sequelae greatly impairing function and likely to make the patient dependent

☐ 1 - Death ☐ Unknown

**Any other comments or information**

______________________________________________

Abbreviations

ASO, antistreptolysin O; AVPU, Alert, Voice, Pain, Unresponsive; CNS, central nervous system; CRP, C-reactive protein; CSF, cerebrospinal fluid; CT, computed tomography; EEG, electroencephalogram; GAS, group A Streptococcus; GCS, Glasgow Coma Scale; iGAS, invasive group A streptococcal disease; IVIG, intravenous immunoglobulin; MRI, magnetic resonance imaging; PANDAS, pediatric autoimmune neuropsychiatric disorders associated with streptococcal infections; PCR, polymerase chain reaction; PICU, pediatric intensive care unit.

### Supplementary Appendix 2. Supplementary tables and figure

#### Supplementary Table 1. Patient characteristics and presentation

| Characteristic | N = 48 |
|---|---|
| **Demographic characteristics** | |
| Median age, months | 57 (20.8-113.3) |
| **Age group, years** | |
| <1 | 3 (6%) |
| 1-4 | 23 (48%) |
| 5-9 | 12 (25%) |
| 10-17 | 10 (21%) |
| **Sex** | |
| Male | 29 (60%) |
| Female | 19 (40%) |
| **Ethnicity** | |
| White | 29 (60%) |
| Asian or Asian British | 2 (4%) |
| Black African, Caribbean, or Black British | 2 (4%) |
| Mixed or multiple ethnic groups | 1 (2%) |
| Other ethnic group | 2 (4%) |
| Unknown | 12 (25%) |
| **Patient source** | |
| Tertiary referral or transfer | 34 (71%) |
| Direct admission | 13 (27%) |
| Outpatient admission | 1 (2%) |
| **Medical background** | |
| Preexisting medical condition[a] | 9 (19%) |
| Prematurity | 3 (6%) |
| Neurological or neurodevelopmental condition | 2 (4%) |
| Cardiac disease | 1 (2%) |
| Diabetes or other endocrine disease | 1 (2%) |
| Kidney disease | 1 (2%) |
| Respiratory disease | 1 (2%) |
| Other | 4 (8%) |
| No preexisting medical condition | 38 (79%) |
| Unknown preexisting-condition status | 1 (2%) |
| Receiving immunosuppressive or immunomodulatory therapy | 0 (0%) |
| **Immunization status** | |
| Fully immunized for age | 33 (69%) |
| Incomplete | 1 (2%) |
| Unknown | 14 (29%) |
| **Risk factors before presentation** | |
| At least 1 neurological or non-neurological risk factor[b] | 38 (79%) |
| **Non-neurological risk factor[b]** | 21 (44%) |
| Pharyngitis or tonsillitis | 12 (25%) |
| Varicella | 3 (6%) |
| Skin or soft-tissue infection | 2 (4%) |
| Other | 7 (15%) |

| Characteristic | N = 48 |
|---|---|
| **Neurological or parameningeal risk factor[b]** | 28 (58%) |
| Otitis media | 10 (21%) |
| Mastoiditis | 10 (21%) |
| Sinus infection | 6 (13%) |
| Head trauma | 5 (10%) |
| Orbital cellulitis | 1 (2%) |
| Ventriculoperitoneal shunt and recent neurosurgery | 1 (2%) |
| Other | 3 (6%) |
| **Clinical presentation** | |
| **Constitutional symptom[b]** | 32 (67%) |
| Fever | 32 (67%) |
| Rash | 5 (10%) |
| Myalgia | 2 (4%) |
| **Neurological symptom[b]** | 41 (85%) |
| Lethargy | 20 (42%) |
| Headache | 16 (33%) |
| Seizures | 14 (29%) |
| Drowsiness | 14 (29%) |
| Focal neurological deficit | 4 (8%) |
| Neck stiffness or pain | 4 (8%) |
| Photophobia | 4 (8%) |
| Coma | 3 (6%) |
| Irritability, behavioral change, or confusion | 3 (6%) |
| Cerebellar signs (ataxia or nystagmus) | 2 (4%) |
| **Ear, nose, throat, or respiratory symptom[b]** | 25 (52%) |
| Earache | 12 (25%) |
| Coryza | 8 (17%) |
| Cough | 7 (15%) |
| Sore throat | 6 (13%) |
| Difficulty breathing | 2 (4%) |
| Epistaxis | 1 (2%) |
| Hearing loss | 1 (2%) |
| Other | 2 (4%) |
| **Gastrointestinal symptom[b]** | 25 (52%) |
| Vomiting | 23 (48%) |
| Diarrhea | 4 (8%) |
| Reduced appetite or oral intake | 2 (4%) |
| **Neurological status on admission** | |
| Glasgow Coma Scale recorded | 29 (60%) |
| **Glasgow Coma Scale score among those recorded[c]** | |
| 15 | 7/29 (24%) |
| 9-14 | 14/29 (48%) |
| ≤8 | 8/29 (28%) |
| AVPU recorded | 12 (25%) |
| Neither Glasgow Coma Scale nor AVPU recorded | 9 (19%) |

*Data are n (%) or median (IQR). Percentages use N = 48 unless another denominator is shown. Read down the left block, then down the right block.*

*[a] Medical conditions were not mutually exclusive. [b] Risk factors, symptom groups and individual symptoms were not mutually exclusive. [c] Glasgow Coma Scale percentages use the 29 children with a recorded score.*

*Abbreviations: AVPU, Alert, Verbal, Pain, Unresponsive; IQR, interquartile range.*

### Supplementary Table 2. Early and late outcomes by CNS phenotype

| CNS phenotype[a] | Children, n (%) | Early death[b] | Early neurological sequelae[b] | Early adverse outcome[b] | Late death[c] | Late death or neurological sequelae[c] |
|---|---|---|---|---|---|---|
| Subdural empyema | 16 (33%) | 1/16 (6%)<br>P = 0.648 | 4/16 (25%)<br>P = 1.000 | 5/16 (31%)<br>P = 0.541 | 1/16 (6%)<br>P = 0.648 | 6/16 (38%)<br>P = 0.547 |
| Meningitis | 13 (27%) | 3/13 (23%)<br>P = 0.121 | 3/13 (23%)<br>P = 1.000 | 6/13 (46%)<br>P = 0.521 | 3/13 (23%)<br>P = 0.326 | 7/13 (54%)<br>P = 0.520 |
| Intracranial thrombosis | 9 (19%) | 0/9 (0%)<br>P = 0.567 | 4/9 (44%)<br>P = 0.237 | 4/9 (44%)<br>P = 0.716 | 0/9 (0%)<br>P = 0.579 | 4/9 (44%)<br>P = 1.000 |
| Extradural empyema | 8 (17%) | 0/8 (0%)<br>P = 0.571 | 1/8 (13%)<br>P = 0.413 | 1/8 (13%)<br>P = 0.130 | 0/8 (0%)<br>P = 0.571 | 1/8 (13%)<br>P = 0.059 |
| Brain abscess | 6 (13%) | 0/6 (0%)<br>P = 1.000 | 3/6 (50%)<br>P = 0.326 | 3/6 (50%)<br>P = 0.662 | 1/6 (17%)<br>P = 1.000 | 4/6 (67%)<br>P = 0.386 |
| Meningoencephalitis | 4 (8%) | 2/4 (50%)<br>P = 0.051 | 2/4 (50%)<br>P = 0.304 | 4/4 (100%)<br>P = 0.017 | 2/4 (50%)<br>P = 0.074 | 4/4 (100%)<br>P = 0.034 |
| Cerebral infarction | 4 (8%) | 0/3 (0%)<br>P = 1.000 | 3/3 (100%)<br>P = 0.018 | 3/3 (100%)<br>P = 0.050 | 0/3 (0%)<br>P = 1.000 | 3/3 (100%)<br>P = 0.082 |
| Hydrocephalus | 3 (6%) | 1/3 (33%)<br>P = 0.292 | 0/3 (0%)<br>P = 0.550 | 1/3 (33%)<br>P = 1.000 | 1/3 (33%)<br>P = 0.343 | 1/3 (33%)<br>P = 1.000 |
| Encephalitis | 1 (2%) | 0/1 (0%)<br>P = 1.000 | 0/1 (0%)<br>P = 1.000 | 0/1 (0%)<br>P = 1.000 | 0/1 (0%)<br>P = 1.000 | 0/1 (0%)<br>P = 1.000 |
| Other | 6 (13%) | 0/6 (0%)<br>P = 1.000 | 1/6 (17%)<br>P = 1.000 | 1/6 (17%)<br>P = 0.384 | 0/6 (0%)<br>P = 1.000 | 1/6 (17%)<br>P = 0.204 |

*Data are n/N (%) for outcomes; cohort percentages use N = 48. Unknown outcomes are excluded from the relevant denominator.*

*[a] Phenotypes were not mutually exclusive. Other phenotypes comprised cerebellitis, ventriculitis/shunt infection, cerebritis (including cerebritis with osteomyelitis) and superior ophthalmic vein thrombosis.*

*[b] Early outcomes were assessed at discharge from the acute treating-hospital admission. Early adverse outcome means death or survival with neurological sequelae.*

*[c] Late outcomes were assessed at the most recent clinical review using the modified Glasgow Outcome Scale. Late death or neurological sequelae includes death or any neurological sequelae at that assessment.*

*P values compare children with versus without each phenotype using two-sided Fisher exact tests. Comparisons are exploratory and unadjusted for multiple testing.*

*Abbreviation: CNS, central nervous system.*

### Supplementary Table 3. GAS detection by specimen source and method

| Specimen source | Culture detected, n (%) | PCR detected[b], n (%) | Culture and PCR[b], n (%) |
|---|---|---|---|
| Pus[a] | 22 (46%) | 5 (10%) | 1 (2%) |
| Blood | 13 (27%) | 3 (6%) | 3 (6%) |
| CSF | 6 (13%) | 4 (8%) | 2 (4%) |
| Throat | 7 (15%) | - | - |
| Ear swab or NPA | 4 (8%) | - | - |
| Skin | 3 (6%) | - | - |
| Other | 3 (6%) | 2 (4%) | 1 (2%) |
| Unknown PCR specimen | - | 1 (2%) | - |

*Data are n (% of the 48-child cohort). More than one specimen source could be recorded per child; column totals may therefore exceed 100%. A dash indicates no detection recorded for that source and method.*

*[a] Pus included surgically drained intracranial or parameningeal collections: subdural or extradural empyema, brain abscess, and mastoid, sinus, orbital or facial collections. Superficial skin specimens were recorded separately.*

*[b] PCR included GAS-specific and broad-range 16S PCR. "Culture and PCR" identifies children with GAS detected by both methods from the same specimen source; these children are included in both method-specific columns.*

*Abbreviations: CSF, cerebrospinal fluid; GAS, group A Streptococcus; NPA, nasopharyngeal aspirate; PCR, polymerase chain reaction.*

### Supplementary Table 4. Blood and CSF markers by CNS phenotype

#### Panel A. Blood inflammatory markers

| CNS phenotype | Children, n (%) | Highest WCC, ×10⁹/L | Highest neutrophil count, ×10⁹/L | Highest platelet count, ×10⁹/L | Highest CRP, mg/L |
|---|---|---|---|---|---|
| Subdural empyema | 16 (33%) | 19.2 (13.2-25.5)<br>n = 16<br>P = 0.759 | 14.2 (7-22.1)<br>n = 16<br>P = 0.718 | 481 (323.2-520.8)<br>n = 16<br>P = 0.063 | 291 (53.8–310)<br>n = 16<br>P = 0.336 |
| Meningitis | 13 (27%) | 16 (12.2-21.4)<br>n = 13<br>P = 0.281 | 13.8 (10.5-19.4)<br>n = 13<br>P = 0.799 | 388 (300-533)<br>n = 13<br>P = 0.026 | 162.4 (85–302.1)<br>n = 13<br>P = 0.693 |
| Intracranial thrombosis | 9 (19%) | 21.3 (15.3-26.9)<br>n = 9<br>P = 0.460 | 14.9 (11.8-23.9)<br>n = 9<br>P = 0.579 | 608 (542-998)<br>n = 9<br>P = 0.051 | 294 (96–332)<br>n = 9<br>P = 0.113 |
| Extradural empyema | 8 (17%) | 15.2 (14.5-16.7)<br>n = 8<br>P = 0.112 | 10.7 (9.1-12.7)<br>n = 8<br>P = 0.146 | 654 (514.2-1027.5)<br>n = 8<br>P = 0.070 | 95.7 (60.5–149.5)<br>n = 8<br>P = 0.135 |
| Brain abscess | 6 (13%) | 19.9 (14.1-24.5)<br>n = 6<br>P = 0.779 | 13.6 (10.1-13.8)<br>n = 6<br>P = 0.342 | 545.5 (456.8-636.5)<br>n = 6<br>P = 0.685 | 12 (5.5–27.3)<br>n = 6<br>P = 0.005 |
| Meningoencephalitis | 4 (8%) | 27.2 (25.3-30.4)<br>n = 4<br>P = 0.065 | 25.1 (22.8-28.7)<br>n = 4<br>P = 0.026 | 580.5 (528.2-675.5)<br>n = 4<br>P = 0.444 | 338.5 (324.5–369.5)<br>n = 4<br>P = 0.022 |
| Cerebral infarction | 4 (8%) | 42.4 (35.3-52)<br>n = 4<br>P = 0.003 | 31 (28.5-33)<br>n = 4<br>P = 0.005 | 838 (554.5-1113)<br>n = 4<br>P = 0.151 | 506.1 (419–555.5)<br>n = 4<br>P = 0.004 |
| Hydrocephalus | 3 (6%) | 25.1 (20.2-32.2)<br>n = 3<br>P = 0.307 | 21.3 (16.6-28.7)<br>n = 3<br>P = 0.250 | 845 (572.5-1022.5)<br>n = 3<br>P = 0.418 | 95.3 (90.2–220.2)<br>n = 3<br>P = 0.766 |
| Encephalitis | 1 (2%) | 24.4 (24.4-24.4)<br>n = 1<br>P = 0.613 | 19.7 (19.7-19.7)<br>n = 1<br>P = 0.516 | 560 (560-560)<br>n = 1<br>P = 0.829 | 75 (75–75)<br>n = 1<br>P = 0.613 |
| Other | 6 (13%) | 17.1 (14.6-20.8)<br>n = 6<br>P = 0.358 | 14.4 (11.9-18.1)<br>n = 6<br>P = 0.720 | 580.5 (496.2-935.5)<br>n = 6<br>P = 0.391 | 85.2 (23.9–251.1)<br>n = 6<br>P = 0.697 |

*Values are median (IQR); n is the number of children with an available numeric result. Cohort percentages use N = 48. NA indicates no available numeric result.*
*P values compare children with versus without each phenotype using Mann-Whitney U tests. Comparisons are exploratory and unadjusted for multiple testing.*
*Phenotypes were not mutually exclusive. Other phenotypes comprised cerebellitis, ventriculitis/shunt infection, cerebritis (including cerebritis with osteomyelitis) and superior ophthalmic vein thrombosis.*
*Abbreviations: CNS, central nervous system; CRP, C-reactive protein; CSF, cerebrospinal fluid; IQR, interquartile range; NA, not available; WCC, white cell count.*

### Supplementary Table 4. Blood and CSF markers by CNS phenotype

#### Panel B. CSF cell counts

| CNS phenotype | Children, n (%) | CSF WCC, ×10⁶/L | CSF lymphocyte count, ×10⁶/L | CSF polymorph count, ×10⁶/L | CSF RBC count, ×10⁶/L |
|---|---|---|---|---|---|
| Subdural empyema | 16 (33%) | 46 (23-2040)<br>n = 3<br>P = 0.670 | 30 (15-821.5)<br>n = 3<br>P = 0.177 | 0 (0-1210)<br>n = 3<br>P = 0.757 | 100 (51.5-2170)<br>n = 3<br>P = 0.554 |
| Meningitis | 13 (27%) | 700 (1-2030)<br>n = 5<br>P = 0.460 | 0 (0-168)<br>n = 5<br>P = 0.616 | 70 (0-532)<br>n = 5<br>P = 0.060 | 132 (100-160)<br>n = 5<br>P = 0.164 |
| Intracranial thrombosis | 9 (19%) | 0.5 (0.2-0.8)<br>n = 2<br>P = 0.164 | 0 (0-0)<br>n = 2<br>P = 0.310 | 0 (0-0)<br>n = 2<br>P = 0.399 | 500.5 (250.8-750.2)<br>n = 2<br>P = 1.000 |
| Extradural empyema | 8 (17%) | 0 (0-0)<br>n = 1<br>P = 0.225 | 0 (0-0)<br>n = 1<br>P = 0.542 | 0 (0-0)<br>n = 1<br>P = 0.624 | 1000 (1000-1000)<br>n = 1<br>P = 0.349 |
| Brain abscess | 6 (13%) | 6 (6-6)<br>n = 2<br>P = 0.921 | 6 (6-6)<br>n = 2<br>P = 0.310 | 0 (0-0)<br>n = 2<br>P = 0.399 | 0 (0-0)<br>n = 2<br>P = 0.038 |
| Meningoencephalitis | 4 (8%) | 2220 (2220-2220)<br>n = 1<br>P = 0.225 | 0 (0-0)<br>n = 1<br>P = 0.542 | 0 (0-0)<br>n = 1<br>P = 0.624 | 21 (2-21)<br>n = 1<br>P = 0.688 |
| Cerebral infarction | 4 (8%) | 1 (1-1)<br>n = 1<br>P = 0.590 | 0 (0-0)<br>n = 1<br>P = 0.542 | 0 (0-0)<br>n = 1<br>P = 0.624 | 1 (1-1)<br>n = 1<br>P = 0.349 |
| Hydrocephalus | 3 (6%) | 2017 (1008.5-3025.5)<br>n = 2<br>P = 0.921 | 806.5 (403.2-1209.8)<br>n = 2<br>P = 0.499 | 1210 (605-1815)<br>n = 2<br>P = 0.399 | 550 (325-775)<br>n = 2<br>P = 0.374 |
| Encephalitis | 1 (2%) | NA | NA | NA | NA |
| Other | 6 (13%) | 7 (0-518)<br>n = 4<br>P = 0.484 | 0 (0-0)<br>n = 4<br>P = 0.094 | 7 (0-28)<br>n = 4<br>P = 0.638 | 116.5 (55.5-370)<br>n = 4<br>P = 0.589 |

*Values are median (IQR); n is the number of children with an available numeric result. Cohort percentages use N = 48. NA indicates no available numeric result.*

*P values compare children with versus without each phenotype using Mann-Whitney U tests. Comparisons are exploratory and unadjusted for multiple testing.*

*Phenotypes were not mutually exclusive. Other phenotypes comprised cerebellitis, ventriculitis/shunt infection, cerebritis (including cerebritis with osteomyelitis) and superior ophthalmic vein thrombosis.*

*Abbreviations: CNS, central nervous system; CSF, cerebrospinal fluid; IQR, interquartile range; NA, not available; RBC, red blood cell; WCC, white cell count.*

### Supplementary Table 4. Blood and CSF markers by CNS phenotype

#### Panel C. CSF biochemistry

| CNS phenotype | Children, n (%) | CSF glucose, mmol/L | Plasma glucose, mmol/L | CSF protein, g/L |
|---|---|---|---|---|
| Subdural empyema | 16 (33%) | 1.5 (1-2)<br>n = 2<br>P = 0.056 | 5.1 (4.8-5.7)<br>n = 3<br>P = 1.000 | 0.6 (0.4–0.7)<br>n = 2<br>P = 0.723 |
| Meningitis | 13 (27%) | 3.1 (2.8-3.1)<br>n = 5<br>P = 0.176 | 5.5 (5-6.3)<br>n = 6<br>P = 0.230 | 0.6 (0.3–0.9)<br>n = 6<br>P = 0.360 |
| Intracranial thrombosis | 9 (19%) | 4.3 (4.3-4.3)<br>n = 1<br>P = 0.331 | 4.7 (4.7-4.7)<br>n = 1<br>P = 0.562 | 0.5 (0.5–0.5)<br>n = 1<br>P = 0.634 |
| Extradural empyema | 8 (17%) | NA | NA | NA |
| Brain abscess | 6 (13%) | 5.2 (5.2-5.2)<br>n = 1<br>P = 0.173 | 5.2 (5.2-5.2)<br>n = 1<br>P = 0.772 | 0.5 (0.5–0.5)<br>n = 1<br>P = 0.874 |
| Meningoencephalitis | 4 (8%) | 2.8 (2.8-2.8)<br>n = 1<br>P = 0.560 | 3.2 (1.6-4.8)<br>n = 2<br>P = 0.914 | 0.5 (0.2–0.7)<br>n = 2<br>P = 1.000 |
| Cerebral infarction | 4 (8%) | 4.3 (4.3-4.3)<br>n = 1<br>P = 0.331 | 4.7 (4.7-4.7)<br>n = 1<br>P = 0.562 | 0.5 (0.5–0.5)<br>n = 1<br>P = 0.634 |
| Hydrocephalus | 3 (6%) | 1.6 (1.1-2.2)<br>n = 2<br>P = 0.106 | 5.8 (5.4-6.1)<br>n = 2<br>P = 0.334 | 0.9 (0.9–0.9)<br>n = 2<br>P = 0.125 |
| Encephalitis | 1 (2%) | NA | NA | NA |
| Other | 6 (13%) | 3 (2.7-3.2)<br>n = 2<br>P = 0.883 | 4.8 (4.6-5.3)<br>n = 3<br>P = 0.712 | 0.2 (0.1–0.3)<br>n = 3<br>P = 0.102 |

*Values are median (IQR); n is the number of children with an available numeric result. Cohort percentages use N = 48. NA indicates no available numeric result.*
*P values compare children with versus without each phenotype using Mann–Whitney U tests. Comparisons are exploratory and unadjusted for multiple testing.*
*Phenotypes were not mutually exclusive. Other phenotypes comprised cerebellitis, ventriculitis/shunt infection, cerebritis (including cerebritis with osteomyelitis) and superior ophthalmic vein thrombosis.*
*Abbreviations: CNS, central nervous system; CSF, cerebrospinal fluid; IQR, interquartile range; NA, not available.*

### Supplementary Table 5. *emm* typing results

| Typing status or *emm* type | n | % |
|---|---|---|
| Performed | 28 | 58 |
| *emm*1.0 | 16 | 57 |
| *emm*6.0 | 2 | 7 |
| *emm*89.0 | 2 | 7 |
| *emm*1.150 | 1 | 4 |
| *emm*3.93 | 1 | 4 |
| *emm*4.0 | 1 | 4 |
| *emm*4.40 | 1 | 4 |
| *emm*6.125 | 1 | 4 |
| *emm*12.122 | 1 | 4 |
| *emm*12.40 | 1 | 4 |
| *emm*66.0 | 1 | 4 |
| Not performed | 15 | 31 |
| Unknown | 5 | 10 |

*Typing-status percentages use the full cohort (N = 48). Percentages for individual emm types use the 28 children who underwent typing. Percentages are rounded and may not total 100%.*

*Abbreviation: emm, gene encoding the streptococcal M protein; emm typing is based on its sequence.*

### Supplementary Table 6. Clinical management and interventions

| Management or intervention | n/N (%) or median (IQR) |
|---|---|
| **PICU and organ support** | |
| PICU admission | 22/48 (46%) |
| PICU stay, days | 3 (2-9) |
| Respiratory support in PICU | 18/22 (82%) |
| Invasive support among children receiving respiratory support | 18/18 (100%) |
| Inotropic support in PICU | 12/22 (55%) |
| **Venous sinus thrombosis and anticoagulation** | |
| Venous sinus thrombosis | 15/48 (31%) |
| Anticoagulation among children with venous sinus thrombosis | 14/15 (93%) |
| Anticoagulation duration, weeks | 14 (12-25.5) |
| **Intravenous immunoglobulin** | |
| IVIG received | 3/48 (6%) |
| Repeat IVIG among children receiving IVIG[a] | 1/3 (33%) |
| **Surgery and procedures** | |
| Surgical intervention | 37/48 (77%) |
| **ENT operations[b]** | **17/37 (46%)** |
| Mastoidectomy | 11/37 (30%) |
| Myringotomy and/or grommet insertion | 5/37 (14%) |
| Functional endoscopic sinus surgery | 3/37 (8%) |
| Other ENT procedure | 2/37 (5%) |
| **Neurosurgical operations[c]** | **25/37 (68%)** |
| Burr hole aspiration | 11/37 (30%) |
| Craniotomy | 10/37 (27%) |
| External ventricular drain insertion | 7/37 (19%) |
| Other neurosurgical procedure[c] | 2/37 (5%) |
| **Other** | **4/37 (11%)** |
| Other procedure[d] | 4/37 (11%) |
| Unknown procedure | 1/37 (3%) |
| Further surgery among children with surgery | 8/37 (22%) |
| Further surgery among children with a neurosurgical procedure | 6/25 (24%) |
| Further neurosurgical procedure among children with a neurosurgical procedure | 4/25 (16%) |

*Data are n/N (%) or median (IQR). Percentages use the denominator shown. Procedure categories were not mutually exclusive.*

*[a] Two children received a cumulative IVIG dose of 2 g/kg and one received 1 g/kg; one child received repeat IVIG.*

*[b] ENT procedures included mastoidectomy, myringotomy and/or grommet insertion, functional endoscopic sinus surgery, and facial or orbital drainage.*

*[c] Neurosurgical procedures included burr hole aspiration, craniotomy, external ventricular drain insertion, epidural drainage, craniectomy and cranioplasty.*

*[d] Other procedures included cardiac, vascular and abdominal procedures, skin debridement/grafting, and antrostomy. Unknown procedures are shown separately.*

*Abbreviations: ENT, ear, nose and throat; IQR, interquartile range; IVIG, intravenous immunoglobulin; PICU, pediatric intensive care unit.*

**Supplementary Figure 1. Factors associated with late outcomes**

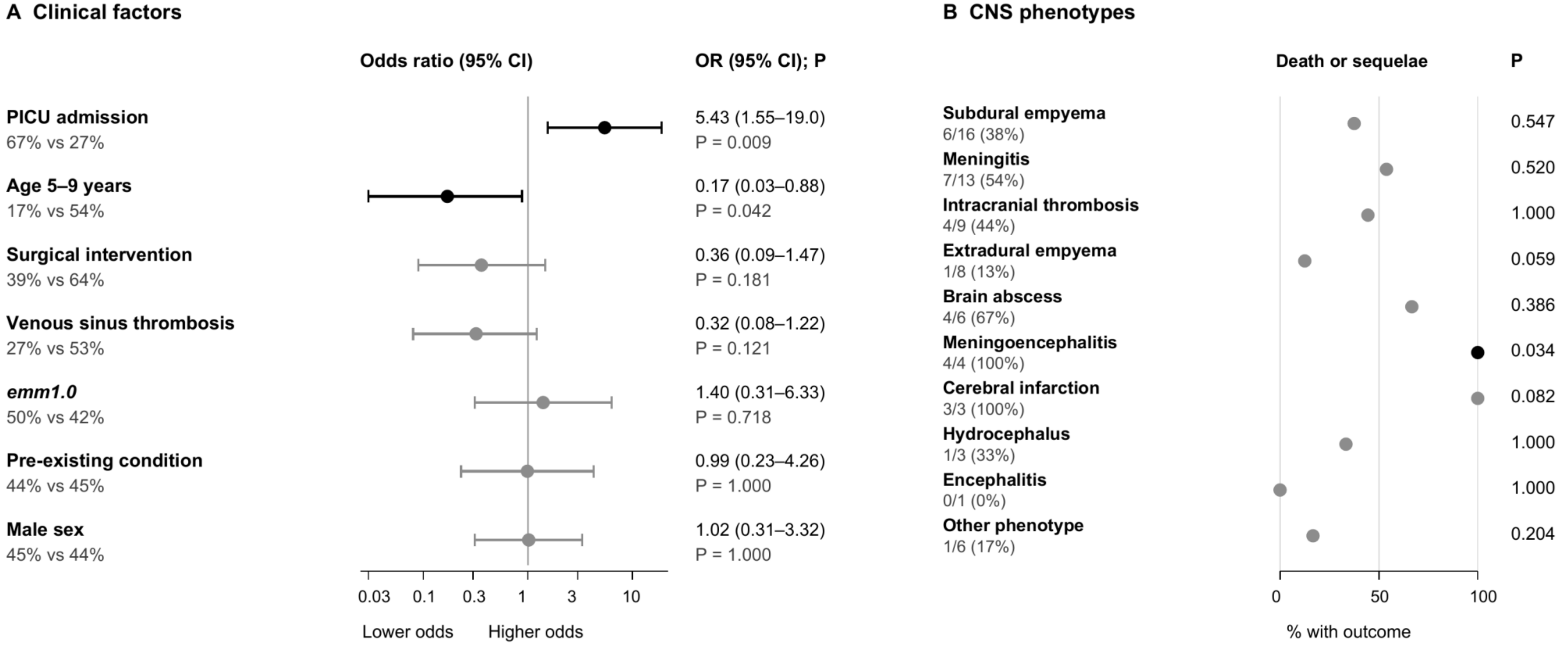


**PICU admission**

*Death: 6/21 (29%) vs 0/26 (0%); P = 0.005.*

*Death or neurological sequelae: 14/21 (67%) vs 7/26 (27%); P = 0.009.*

**Cerebral infarction**

*Neurological sequelae: 3/3 (100%) vs 12/44 (27%); P = 0.028.*

*One phenotype case had an unknown late outcome.*

**Meningoencephalitis**

*Death or neurological sequelae: 4/4 (100%) vs 17/43 (40%); P = 0.034.*

**Age 5-9 years**

*Full recovery: 10/12 (83%) vs 16/35 (46%); P = 0.042.*

*Late outcomes were assessed at the most recent clinical review; 47 children had known outcomes. Death or neurological sequelae corresponds to modified Glasgow Outcome Scale scores 1–4.*

*A: Circles show odds ratios and horizontal lines show 95% CIs on a logarithmic scale; the reference line is OR = 1. Percentages compare children with versus without each factor; age 5–9 years is compared with all other ages, and emm1.0 with other typed strains. B: Circles show the proportion with late death or neurological sequelae; labels give n/N (%). Phenotypes were not mutually exclusive.*

*P values are from two-sided Fisher exact tests, without adjustment for multiple comparisons. Black symbols indicate P < 0.05; grey symbols indicate P ≥ 0.05. Unknown outcomes are excluded.*

*Abbreviations: CI, confidence interval; CNS, central nervous system; emm, M-protein gene sequence type; OR, odds ratio; PICU, pediatric intensive care unit.*
